# Prevalence, risks and outcomes of frailty in People Experiencing Homelessness: a protocol for secondary analysis of Health Needs Audit data

**DOI:** 10.1101/2023.12.08.23299735

**Authors:** Jo Dawes, Kate Walters, Rachael Frost, Emmanouil Bagkeris, Alexandra Burton, Debra Herzberg, Andrew Hayward

**Author notes:** Corresponding author (JD).

## Abstract

Frailty describes a health state related to ageing where people become less resilient to health challenges and more likely to have adverse outcomes if they become unwell. People experiencing homelessness (PEH) are known to have poor health, with research suggesting that many become frail at a younger age than the general population. Previous research using small-scale primary data collection suggests that the prevalence of frailty in homeless populations varies widely (16-55%), with variations in sample sizes and settings partially accounting for differences in current estimates. The prevalence, risks, and outcomes of frailty in PEH are poorly understood. We propose to carry out a secondary analysis of existing health survey data collected from 2,792 PEH. This will involve creating a Frailty Index (FI) to identify frail people within the dataset. Regression analyses will be used to identify associations between potential risk factors and outcomes of frailty in this population. This protocol will: 1) Outline the creation of a FI to assess the frailty prevalence within a dataset of health information collected from a cohort of PEH and 2) Describe proposed methods of regression analysis for identification of associations between frailty and risks factors/outcomes of frailty in the cohort of PEH within the dataset. The processes described in this paper can inform future development of FIs in other datasets. It is expected that the FI created will be an appropriate and robust method for identifying frailty in a cohort of PEH and results of the secondary data analysis will provide a more robust estimate of the associations between frailty and risk factors/outcomes.

## Introduction

### Background

Frailty is a health state related to aging where multiple body systems gradually lose their reserves and resilience, which increase risks of adverse outcomes (e.g. prolonged recovery time, disability or death) and health service usage (e.g. hospitalisation, attendance to accident and emergency or greater primary care usage).(1) There are two theoretical approaches to assessing frailty. One is the phenotypic model(2) where frailty is thought to be represented by the presence of up to five characteristics of frailty (unintended weight loss, reduced muscle strength, reduced gait speed, self-reported exhaustion and low energy expenditure). The alternative model is that of cumulative deficit(3) whereby as a person accumulates health deficits (e.g. loss of hearing, low mood, falls, various diseases) which can happen as they age, the deficits combined then form a “frailty index”. It is the model of cumulative deficit that is applied in this research protocol, with the FI recognised as strongly associated with adverse outcomes including mortality and disability.(4)

Homelessness can be considered as a continuum, at one extreme defining people experiencing homelessness (PEH) solely by their absence of shelter, at the other extreme, is the inclusive definition that a PEH can be someone without access to shelter that meets the basic criteria considered essential for health and social development. For the purpose of this study, PEH are defined as fitting the FEANTSA definitions of “roofless” (e.g. no fixed abode, living in a public space) and “houseless” (e.g. living in hostel, refuge, shelter, temporary accommodation).(5)

PEH have substantially poorer health than the general population, with cohort studies showing 3-6 fold increased mortality risks, high levels of chronic illness and mental health problems.(6, 7) In the general population of England, 8.1 [95% CI 7.3-8.3]% of people aged >50 years are estimated to be frail.(8) Research suggests prevalence of frailty amongst PEH ranges between 16% and 55%,(9–11) often presenting at a younger age. However, frailty in PEH is poorly understood, with studies to date exploring it using primary data collection in specific settings, such as a single homeless hostel, and with relatively small sample populations (range n=33-247).(9, 10) PEH are known to have a high need for and usage of healthcare, which frailty may contribute to, so it is important to identify frailty in this population. To build on the existing literature, this research proposes to explore frailty using secondary analysis of a dataset of health needs information collected from PEH. This will allow for inferences to be made from a far larger sample than previous research has achieved. Moreover, within this dataset, a broad spectrum of types of homelessness is represented, thus allowing for more accurate assessment of prevalence of frailty and associated risk or outcomes in PEH and therefore more generalisable results.

### Research questions

This protocol describes the plan for a cross-sectional analysis of survey data that will address the following research questions:

- What is the prevalence of frailty amongst PEH in England?
- What risk factors and/ or outcomes are associated with frailty in this this population?

### Hypotheses being tested

The Homeless Link dataset contains a broad spectrum of data variables which could be explored for associations with frailty. To avoid data dredging(12) we have conducted preliminary exploration of potential variables with a panel of experts (including researchers and academics with experience of frailty and FI development, medical doctors working in general practice, elderly care, drug and alcohol services, homelessness or frailty) familiar with frailty and/ or homelessness and patient and public involvement and engagement activities (PPIE) with PEH and the staff who care for them to explore, discuss and further refine our hypotheses. Table 1 outlines the hypotheses that will be tested:

**Table 1:** Hypotheses and null hypotheses to be tested by the proposed research.

|  | Hypothesis | Null hypothesis |
| --- | --- | --- |
| **Prevalence of frailty in PEH** | Prevalence of frailty in PEH in England is greater than prevalence reported in the general population (based on comparative literature) | Prevalence of frailty in PEH in England is the same as the prevalence reported in the general population (based on comparative literature) |
|  | The mean age of PEH identified as frail in England is younger than the mean age of people identified as frail in the general population (based on comparative literature) | The mean age of PEH identified as frail in England is the same as the mean age of people identified as frail in the general population (based on comparative literature) |
| **Risks factors of frailty in PEH** | Frailty in PEH is positively associated with addiction (e.g. problematic drug or alcohol use) | There is no association between frailty and addiction (drugs or alcohol) in PEH |
|  | Frailty in PEH is positively associated with impaired cognitive development (e.g. learning disability or autistic spectrum disorders) | There is no association between frailty and impaired cognitive development (e.g. learning disability or autistic spectrum disorders) |
|  | Frailty in PEH is positively associated with sociodemographic factors (e.g. prison, sex work, childhood experience of the care system) | There is no association between frailty and sociodemographic factors (e.g. prison, sex work, childhood experience of the care system) |
| **Outcomes of Frailty in PEH** | Frailty in PEH is positively associated with health care usage (e.g. hospital admission, ambulance, accident and emergency use and General practice) | There is no association between frailty and health care usage (e.g. hospital admission, ambulance, accident and emergency use and General practice) |
|  | Frailty in PEH is negatively associated with preventative health care (e.g. vaccination) | There is no association between frailty and preventative health care (e.g. vaccination) |

### Aims and contribution

The aim of this research is to use an existing dataset of health needs audit data, collected from PEH using a one-to-one, interviewer-led survey, to calculate frailty prevalence and identify frailty risk factors and outcomes of frailty in that population. This study will contribute in two ways. Firstly, it will provide a calculation of prevalence of frailty in this population from a far larger dataset than any previous research has explored-thus improving accuracy and precision of this calculation with the potential for supporting investment in services to address frailty in PEH. Secondly, it will explore risk factors for and outcomes of frailty, potentially supporting targeting of interventions to those most at risk of frailty or of poor outcomes of frailty. The intention is to provide information that can help direct resources to prevent and address frailty in PEH rather than to determine causality.

## Materials and Methods

### Design and setting

The design of this study is an analytical, cross-sectional study, using secondary analysis of health needs audit survey data collected from PEH. The primary dataset was collected from PEH in England between 2015 and 2022. Settings for data collection included homeless accommodation services, outreach, day centres, night shelters, and specialist support services.(13)

### Dataset

The data comprises health information (demographics, physical health, mental health, drug and alcohol use, health service usage, wellbeing and preventative healthcare)(13) collected by a partnership of service providers, recruited by the audit commissioner, homelessness charity Homeless Link. It is important to note that although this survey tool contains substantial health information relevant to frailty, it was not designed with frailty assessments in mind. The data were collected between 2015 and 2022 from people across England who were either homeless at the time of the survey or had experienced homelessness in the preceding 12 months. Data were collected in accordance with GDPR (2018) by trained individuals using the Homeless Health Needs Audit (HHNA), an interviewer administered health survey tool. The advantage of using this dataset is that all participants were either currently homeless or had recently experienced homelessness leading up to the survey. This is important because there are substantial problems with identifying PEH from other, more commonly used national datasets of health information. For example, PEH may be difficult to identify due to non-disclosure of homelessness status, providing an address of a relative/ friend, or stating a hostel or temporary accommodation as their address. To ensure the research team do not have any prior knowledge of any patterns or summary statistics of the data at this stage of planning, no reports of information relating to frailty have been shared with the research team prior to study design and protocol registration. Data was accessed by the UCL research team on 25^th^ July 2023. Data was sufficiently annonymised that no information which made individual participants identifiable could be accessed.

### Sample size, inclusion, and exclusion criteria of the HHNA dataset

The sample contains data from 2,792 participants who have consented to their anonymous data being collected, analysed, and reported. Inclusion criteria are:

- Participant living in England
- Participant homeless or experienced homelessness within preceding 12 months of survey
- Participant provided survey responses to at least 20/ 43 (46.5%) survey questions

Sample size is determined by the number of people surveyed using the HHNA tool and who have provided their consent for their anonymous data to be used for research purposes. The data were collected before the current protocol was conceived, so the sample size can only be based on the available data.

### Participants and recruitment

Oganisations supporting PEH in England partnered with Homeless Link to collect HHNA data. This involved training partner organisation staff in how to administer the HHNA survey tool in their local area. The recruiting organisations had detailed understanding of the services and landscape of homelessness in their local area, so could target and reach areas when PEH would be, thus optimising opportunities for anyone eligible to participate to be surveyed. Anyone experiencing homelessness within 12 months of the time of interview was invited to participate in the HHNA survey. Informed consent was obtained by providing potential participants with an information sheet outlining the study and how their data would be used. All participants agreeing to be surveyed provided written consent to participate and for their data to be analysed and reported on. Surveys took approximately 30 minutes to complete and were usually undertaken in 1:1 case work sessions.(13)

### How materials will be selected and used

For the purposes of research planning, Homeless Link shared the HHNA survey tool with our research team, allowing for creation of a variables and response list (supplementary file 1). Although Homeless Link descriptive reports of HHNA data are published in the public domain, no secondary analysis has previously been carried out in this cohort relating to frailty.

A Data Sharing Agreement was created between Homeless Link (Data Providers) and UCL (Research Project lead) prior to sharing of data. Also, to comply with the consent provided by participants, all researchers who will access the data were awarded honorary contracts of employment by Homeless Link. The dataset was anonymised by Homeless Link before it was shared with UCL, so no names, unique identification numbers, participants’ dates of birth or any other personally identifying information was shared. Data will be used to summarise the characteristics of the survey population, create a frailty index, calculate the prevalence of frailty, and identify associations between risk and outcome variables and frailty. The HHNA dataset has been shared with the research team in its raw data form, with cleaning currently being undertaken.

### Research processes

The dataset will be cleaned to remove variables irrelevant to the scope of this study. Anomalies in categorisation will be identified and incorrect or incorrectly formatted data will be removed or corrected.

Descriptive statistics will be used to summarise the characteristics of the sample population, including: Age, gender, sexual orientation, ethnicity, immigration status, employment, history of prison, domestic violence, being in the armed forces, local authority care, disability, housing status, mental health conditions, cognitive condition, alcohol consumption, drug use, smoking, GP registration, average meals per day consumed, perceived health, physical health conditions and healthcare utilisation.

### Creation of Frailty Index

Procedures for creating a FI for datasets is outlined by Searle et al(14) and Theou et al(15) who describe step-by-step procedures for FI creation. It is recommended that inclusion of variables in an FI, is based on the following criteria:(14, 15)

1. Variable must be health-related(14, 15)
2. Variable must increase with age(14, 15)
3. Variable must not saturate too early (e.g. not be universal in the adult population by midlife)(14)
4. The total included variables must cover a range of physiological systems and processes.(14, 15)
5. Variable must be present in at least 1% of study population(14, 15)
6. Variable must have no more than 5% missing data(14, 15)
7. Highly correlated screen variables (r>0.95) will be excluded by removing the variable with the lowest response rate (15)

### Modified Delphi for FI creation

In recognition that there is a degree of subjectivity regarding which variables from the HHNA dataset should/ should not be included in the FI, a modified Delphi process was adopted for creation of the FI. The Delphi method is a structured method of developing consensus amongst a panel of experts and is an accepted approach for this purpose.(16) A panel of ten experts working in the field of frailty and/or inclusion health in the UK or Canada were invited by the research team to contribute (six agreed to participate). All who agreed to participate were then invited via email, then met online for a preliminary meeting to determine the nature of their contribution, including agreeing how they would inform the selection of variables. The adapted Delphi method is outlined in Fig 1.

**Fig 1:**
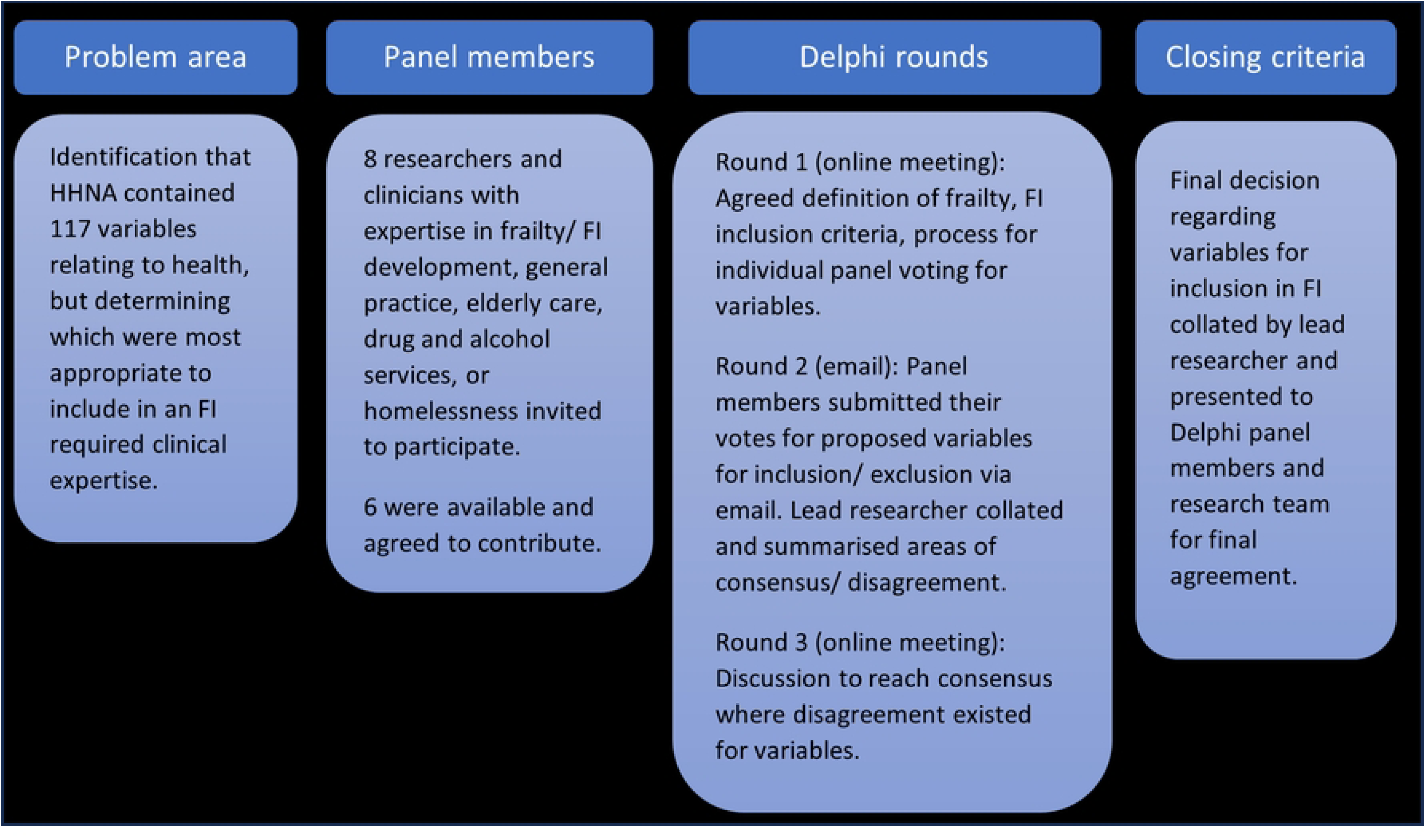
Outline of modified Delphi Method for identification of variables.

A spread sheet of all 117 potential variables was circulated amongst the expert panel, with each member independently reviewing the variables and stating on the spreadsheet whether each variable should be included in either the FI, based on the stated inclusion criteria (e.g. whether they are health related; increase in prevalence with age; and whether they did not saturate early) or considered a risk factor/ outcome for the secondary data analysis. A deadline was set for return of completed spreadsheets to the lead author (JD), whereupon she collated responses, identifying areas of consensus and disagreement. A second meeting was convened to share the findings of the first round and to discuss/ agree any differences of opinion with the panel. After two Delphi rounds with the expert panel, of the potential 117 variables in the HHNA, 41 were viewed as potentially viable for inclusion in a FI for this dataset. Searle et al suggest that 30-40 variables are optimal.

After the dataset is cleaned and reviewed, missing data will be identified, the reason for missingness considered and how missing data may impact inclusion of each variable in the FI and data analysis. Following completion of data scoping, the lead author will determine if the proposed variables cover a range of physiological systems and processes. The final variable list will be circulated with the expert panel, providing an opportunity to review the FI as a whole and comment on its appropriateness to identify frailty in a homeless cohort, including any concerns they have about its likely accuracy.

### Determining frailty using FI

To calculate each participant’s FI score, the sum of the deficit scores will be divided by the total number of deficits measured:

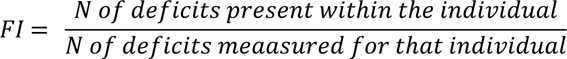

Although any FI score ranges from 0 to a theoretical maximum of 1, it is noted that they consistently show a sub-maximal limit at around 2/3 of the deficits considered. For example, if a frailty index is composed of 60 items, the most items that any participant will likely indicate as being relevant to them is up to 40.(14, 17) For the purposes of analysis, there is debate regarding the determination of a cut off value of a dichotomous “frail” or “not frail”,(18) whereas keeping the FI sores as a continuous variable, allows for analysis to determine how frailty scores change with associated risk factors or outcomes. However, for identifying prevalence of frailty, a cut off figure is required, with different studies citing different values. A systematic review of prevalence of frailty in community-dwelling older people described a FI with a cut off of 0.3.(19) Whereas Rockwood et al suggest 0.25 is generally considered of value.(20) The Fried Phenotypic model classifies people as “not frail”, “pre-frail” and “frail”.(2) To reflect these subgroups, Song et al(21) proposed scores of FI ≤ 0.08 as ‘non-frail’, FI ≥ 0.25 as ‘frail’, and the rest as ‘pre-frail’.(21) For the purposes of this research we will follow the work of Song et al(21) to determine non-frail, pre-frail and frail people in the dataset, since we believe the inclusion of a pre-frail category is likely to be useful in a cohort of PEH, whose age range is younger than many groups where frailty has been researched (the predominant age range is 25-54 years in the HHNA dataset). This decision is supported by the work of Gordon et al(18) whose systematic scoping review indicated 0.25 as the most commonly used score to determine frailty.

### Coding individual variables in FI

All binary variables will be coded using ‘0’ for no deficit and ‘1’ for presence of a deficit. Ordinal and continuous variables will be accommodated by grading the continuum into a rank or score between 0 (where there is no deficit) and 1 (where there is a deficit). For example, a continuous score (such as, rate your health out of 100, where 100 is best health and 0 is worst health) a score of 100 will be rated 0, and a score of 50 would be rated 0.5 etc.

### Analysis Plan for identification of associated risks and outcomes of frailty

#### Public and Patient Involvement and Engagement (PPIE)

In recognition that people who are affected by research should have a say in how it is designed, undertaken and disseminated,(22) people with lived experience of homelessness and frailty and those involved in their care, have contributed to a series of PPIE activities informing this research to date. Firstly, in June 2021, an online session was run in partnership with “Expert Focus”, an organisation that supports PEH to engage with research activities, by minimising the impact of digital exclusion. During the session we discuss conceptual ideas for the research proposal and funding application and the attendees provided feedback on our Plain English Summary of the research, which had been circulated in advance.

After successfully securing of research funding, two further workshops were carried out in November 2022 (one with PEH and one with clinicians working with PEH) which allowed for more detailed consideration of this research. A recommendation from PEH that emerged addressed the importance of language of frailty, with attendees suggesting that using words such as “wellbeing” or “general health” might be more effective for communicating research with PEH, stating that the term “frailty” carried unappealing connotations of weakness and illness that PEH may not wish to identify with.

Finally, review of the survey tool and dataset highlighted many variables which could be considered appropriate for exploration of associations with frailty. It is considered poor research practice to simply explore all potential variables for associations, as this risks generating misleading results.(12) So, in October and November 2023, to understand which variables should be prioritised for statistical exploration of association with frailty, two PPIE workshops (one in person and one via video call) were held with health and social care outreach clinicians working with frail PEH. Additionally, a day of outreach PPIE was held with PEH who themselves were frail to discuss these. Through informal and accessible discussions about the key areas of health in the HHNA, the research team were able to refine their hypotheses and variables of interest.

### Statistical models

Descriptive statistics (proportions, means, standard deviations, medians and interquartile ranges depending on whether a variable is categorical or continuous numeric—normally distributed or skewed) will be used to summarise the population characteristics and state the prevalence (proportion of sample population) of frailty and pre-frailty in the cohort. The creation of an FI for this data allows for it to be expressed both continuously and categorically (not frail, pre-frail, frail). Univariable and multivariable logistic regression analysis will be used to assess the factors associated with frailty when describing associations with frailty expressed categorically and linear regression will be used when expressing frailty on a continuous scale. Regression coefficients and 95% confidence intervals will be reported. In the cases of some variables, it might be debatable whether they are a risk factor, an outcome or risk factor and/or an outcome of frailty. For example, if a relationship between alcohol or drug use and frailty is identified, the cross-sectional nature of this research means it will not be possible to determine the direction of that relationship, i.e., does increased alcohol or drug use issues result in increased frailty? Or does the presence of frailty cause someone’s alcohol or drug use to be altered? Directional acyclic graphs (DAGs) will be used to identify potential confounding factors that should be accounted for in the multivariable models designed to test associations between different relevant variables and frailty amongst PEH.

### Transformations

Data transformation is a technical statistical process that can be used to make data better meet distributional assumptions for parametric tests. It is expected that not all variable data will be normally distributed, so transformations may be required. Transformations will be guided using the Tukey’s Transformation Ladder.(23)

### Inference criteria

Linear and logistic regression analysis will be used to identify associations between dependent and independent variables. In the case of risk factor analysis (for example past adverse experiences) the exposure would be the independent variable and frailty would be the dependent variable.

### Data exclusion

Assessment for exclusion of variables was carried out based on the most recent iteration of the survey tool. Prior to receiving the dataset, it was possible to identify 14 variables that would not be useful to this research, such as variables with limited responses that only applied to participants limited by sex and age range (e.g. uptake of cervical smear, uptake of mammogram, access to free sanitary products) and therefore were not shared by Homeless Link (supplementary file 2). Following creation of the FI a further 22 variables were identified through the PPIE process with clinicians that were not relevant to risk factor or outcome analysis, and so could be excluded. Once the dataset is cleaned, we will identify if any further data needs to be excluded. For example, some variables within the HHNA were only collected in more recent iterations of the survey tool. In these cases, where there are substantial levels of missing data, the variables may need to be excluded from the frailty index and/or the regression analysis.

### Missing data

Once the dataset is cleaned and basic summarises of variables and observations made, we will gauge the volume and patterns of missing data. Where possible, we will determine the likely reason for missing data (completely missing at random, missing at random or missing not at random) by observing patterns in the missingness and communicating with our colleagues at Homeless Link who have detailed insights into the data collection processes. Data might be missing at random if some data collectors failed to ask all questions on the survey tool, or missing not at random if some variable questions do not exist in survey waves or responses to contentious topics, such as sexuality or connection to criminal activity. There are established strategies to deal with missing data. For example, with variables selected to be in the FI, it may be possible to address missing data by coding the variable to binary, with 1= known to have condition x and 0= not known to have condition x, where 0 will also be the code for missing data. However, a limitation of this could be an underestimation of prevalence of conditions contributing to the FI, thus potentially underestimating levels of frailty in the cohort. After identifying all complete cases from the dataset, we will determine if multiple imputation, complete case analysis or coding “missing” as a variable category, may be an appropriate strategy depending on the extent, patterns, and causes of missingness.

### Exploratory analysis

Exploratory descriptive statistics (frequency, calculation of means and Standard Deviations) will be reported to show the characteristics of the participant sample and identify if any of the variables are sufficiently uncommon within the dataset (>1%) or have >5% missing data, leading to removal from the FI. When the FI is finalised, prevalence of frailty in the population will be calculated by selecting all participants with a minimum of 80% complete observation data (Theou et al(15) state that a frailty index score should not be calculated for individuals missing more than 20% of the frailty index items) and determining their individual frailty score. Once calculated, the frailty scores for each individual participant will become an additional continuous variable that can be used, to carry out linear regression to explore the associations between frailty in PEH and risks and outcomes.

### Data management plans

The data comprises anonymised human health information, gathered from people who were homeless at or within 12 months of the data being collected. Data is currently securely stored and processed by Homeless Link (data owner) and UCL (research group, data controller and processor). A Data Sharing Agreement (DSA) between Homeless Link and UCL is place. The data is stored in the UCL Data Safe Haven (DSH), a secure server suitable for the safe storage and analysis of healthcare data. The dataset contains sensitive data (special category data, e.g. ethnicity, health and sex life or sexual orientation)(24) however, the data were anonymised by Homeless Link before transfer to UCL, so the risk of deductive identification of any individual is extremely low. The UCL policy on retention and deletion of data states that data must be stored for 10 years following completion of the project (e.g., publication of final report). After this point the data will be deleted from the UCL DSH.(25)

### Ethical considerations and declarations

Ethical approval for this study was provided by the Research Ethics Committee, University College, London [Project ID: 25071/001] on 7 June 2023. This secondary analysis of existing data, originally collected by a partnership of homelessness service providers recruited by the audit commissioner, Homeless Link, then anonymised by Homeless Link prior to sharing with UCL carries very low risk ethically. Because the data has already been collected, it carries no additional time burden or cost to participants. There is no conflict of interest between the research team, Homeless Link or the participants who have provided data for the research. The presence of a robust DSA agreement between Homeless Link and UCL minimises the risk of data breaches. Additionally, in the reporting of research results, all findings will be reported using a minimum cell size of 10, with specific identifiers, such as age bands location kept broad in reporting.

When balancing the risks and benefits of this research the benefits far outweigh the risks. Although there is unlikely to be direct benefit of this research to any individual who provided their data, the benefits in the long term could highlight a health need in this otherwise vulnerable population and inform policy, practice, investment in services and targeting of services to improve the health landscape for PEH.

### Status and timeline of study

At the point of protocol submission, all ethical approvals, data sharing agreements and data transfer are complete. Data review and cleaning are currently underway. For frailty index creation, the modified Delphi process with the expert panel is complete and a preliminary proposed list of variables for inclusion exists. The next phase for the FI will involve screening of these variables in line with recommendations of Searle et al(14) and Theou et al(15) to exclude any proposed variables which do not meet the inclusion criteria. PPIE activities are underway to prioritise variables for exploration of risk and outcomes of frailty. Data analysis to report prevalence of frailty and risk factor/ outcome associations is expected to be complete by May 2024.

## Discussion

A strength of this proposed research is that it will provide a new level of understanding of frailty amongst PEH in England. By using a large dataset of healthcare data collected entirely from people who were homeless within a 12-month period prior to undertaking the survey, this study enables accurate representation of the population on a far larger scale than has been possible in previous small-scale studies exploring frailty. Another strength is the robust approach to development of the Frailty Index. By using an established framework for considering which variables should contribute to frailty indexes and an adapted Delphi method, undertaken in a systematic way and by inviting input from clinical and academic experts in the fields of frailty and homelessness, the development of the FI is well informed. Finally, involving, and engaging people with lived experience of homelessness and being frail and the staff who care for them in prioritising which variables should be considered for exploration of associations with frailty and stating hypotheses in advance of data analysis means that the risk of type one errors and poor statistical practices, such as data dredging are removed. Limitations of this research must also be acknowledged. Firstly, the development of the FI for this research can only be applied to the Homeless Link dataset. So, although the principles of how the FI is developed can be mirrored by other researchers, the FI itself cannot be transferred to a different database which has not collected the variables included in the index. However, it will be made public and shared, so any future research related to frailty and using the Homeless Link database or other databases with relevant data, can harness the knowledge gained from this proposal and benefit future research. Also, this cross-sectional observational study does not have a comparison group for the assessment of prevalence of frailty, so we will make comparisons with findings of prevalence in the literature-recognising that this will likely have differences in population characteristics and how frailty was determined. These limitations will be noted in reporting. Additionally, we cannot identify temporal associations of risk factors and outcomes with frailty status, thus it is not possible to make any interpretations of the findings related to causality. It must be noted that due to data collection and anonymisation processes, it is not possible to identify repeat observations from the dataset, for example, if participants have completed the HHNA survey on more than one occasion, or how that may impact analysis. Amendments to the proposed study will be dealt with appropriately depending on the nature of the amendment. For example, amendments related to ethics will be submitted to the UCL Research Ethics Committee.

This research will have a broad dissemination plan, which will be widely accessible to a variety of audiences. We intend for PEH to learn about this research and/or engage with it in the future, by publicising it in an accessibly written format in a nationally published magazine widely viewed by PEH, such as Pavement or the Big Issue. We also intend to reach a wide range of staff working in the field of inclusion health, including those involved in commissioning of inclusion health services (e.g. Integrated Care Boards) or policy regarding PEH (e.g. OHID or UKHSA or DHSC or DLUCH) by communicating through peer reviewed publications, relevant conferences, staff training and webinars. We anticipate that there will be a degree of snowballing with this approach-as staff working in Inclusion Health learn about our findings, in turn it should reach many PEH through communication in their places of accommodation, support services or healthcare. We do not anticipate any ethical issues to arise from this. By sharing our research there are important implications, including a greater understanding of the issue of frailty in PEH a topic which is widely acknowledged as important within the community of those with lived experience of homelessness and people who care for them. By quantifying the prevalence of frailty, we create an objective advocacy tool for policy change and by identifying associations with frailty, we gain insights into how interventions and future research should be targeted to address this important health issue.

## Authors’ contributions

JD, KW, RF and AH were responsible for design, methodology and conception of this research protocol. DH was responsible for data collection and data management. JD and AH were responsible for liaison with colleagues at Homeless Link to acquire the data for analysis. JD was responsible for funding acquisition, project administration, drafting and critically revising the article for important intellectual content. AH, KW, EB, RF and AB are all responsible for supervision of this research. All authors have approved the final version to be published and agree to be accountable for all aspects of the work in ensuring that questions related to the accuracy or integrity of any part of the work are appropriately investigated and resolved.

## Metadata

### Funding

JD is funded by a National Institute of Healthcare Research (NIHR) Doctoral Research Fellowship [NIHR302285]. The funder’s role in this research is to provide full fellowship funding for the lead researcher (JD), as part of a wider research training programme and completion of PhD.

### Data availability

This study used third party data made available under licence that the author does not have permission to share. Requests to access the data should be directed to Homeless Link at: www.homeless.org.uk or.

### Associated content

STROBE checklist

## Acknowledgements

The authors would like to thank Sophie Boobis from Homeless Link for her commitment, time and support to this proposed research. We would also like to thank our PPIE and Delphi expert panel contributors for sharing their knowledge and expertise in the development of this research.

## Supporting information

S1: Supplementary file 1: All variables collected using Homeless Link HHNA

S2: Fig 1: Outline of modified Delphi Method for identification of variables

S3: Supplementary file 2: HHNA variables excluded and not shared following survey tool review

S4: STROBE checklist

